# A co-infection model for Two-Strain Malaria and Cholera with Optimal Control

**DOI:** 10.1101/2020.08.18.20177329

**Authors:** K. U. Egeonu, S. C. Inyama, A. Omame

## Abstract

A mathematical model for two strains of Malaria and Cholera with optimal control is studied and analyzed to assess the impact of treatment controls in reducing the burden of the diseases in a population, in the presence of malaria drug resistance. The model is shown to exhibit the dynamical property of backward bifurcation when the associated reproduction number is less than unity. The global asymptotic stability of the disease-free equilibrium of the model is proven not to exist. The necessary conditions for the existence of optimal control and the optimality system for the model is established using the Pontryagin’s Maximum Principle. Numerical simulations of the optimal control model reveal that malaria drug resistance can greatly influence the co-infection cases averted, even in the presence of treatment controls for co-infected individuals.

## 1 Introduction

Malaria is one of the febrile illnesses and the most common deadly disease in the world caused by one or more species of plasmodium. These are *Plasmodium falciparum*, *Plasmodium vivax, Plasmodium ovale*, *Plasmodium malariae*, and *Plasmodium knowlesi*. Approximately half of the world population is at risk of malaria. Most of malaria cases and deaths occur in sub-Saharan Africa [1]. According to the World Malaria Report 2018, there were about 228 million cases of malaria and an estimated 405 000 deaths in 2018 [2]. More than 50% of all cases were reported in six countries in Africa, with Nigeria recording 25% of the total global cases. Other African countries include Democratic Republic of Congo (12%); Uganda (5%); Cote d’Ivoire, Mozambique and Niger (4% each) [2].

Cholera is an acute, diarrheal disease caused by infection of the intestine with the toxigenic bacterium *Vibrio cholerae serogroup O1 or O139*. About 2.9 million persons are estimated to be infected worldwide, with roughly 95,000 deaths annually [3]. Mostly, the infection is mild or without symptoms, although can sometimes be severe. “Approximately 10% of those infected with the cholera will develop severe disease characterized by profuse watery diarrhea, vomiting, and leg cramps. In these people, rapid loss of body fluids leads to dehydration and shock. Without treatment, death can occur within hours” [3].

Mathematical models have been used extensively in studying the behaviour of infectious diseases [4, 5, 6, 7, 8, 9, 10, 11]. A lot of models have been developed for the dynamics of the co-infections of two diseases [12, 13, 14, 15, 16, 17, 18]. In this study, we shall be investigating the impact of malaria drug resistance on the co-infection malaria and cholera, using a mathematical model. Furthermore, optimal control analysis shall be carried out on the model to assess the impact of some control strategies.

## 2 Model formulation

The total human population at time *t*, denoted by *N*_H_(*t*), is divided into eleven mutually exclusive classes, namely: susceptible humans (*S*_H_(*t*)), untreated individuals with the sensitive malaria strain 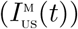, individuals treated of sensitive malaria strain 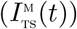, infectious individuals with resistant malaria strain 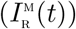, individual that recovered from malaria infection *R*_M_(*t*)), indivdiduals with cholera infection (*I*_C_(*t*)), individuals who have recovered from cholera (*R*_C_(*t*)), individuals untreated of sensitive malaria and infected with cholera, 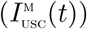, infectious individual treated of sensitive malaria but with cholera infection 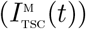, individual with drug resistant malaria and cholera 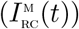, individuals who have recovered from both malaria and cholera (*R*_MC_(*t*)). Thus

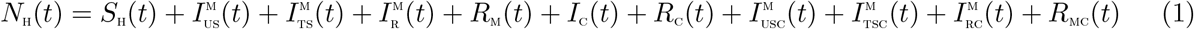

The total vector population *N*_V_ is divided int three mutually exclusive classes: susceptible vectors, *S*_V_, vectors infected with the sensitive malaria strain, *I*_VS_, vectors infected with the resistant malaria strain *I*_VR_. Thus, *N*_V_ = *S*_V_ + *I*_VS_ + *I*_VR_. Also, the bacteria population is given by (*B*_C_). Hence, the two strain malaria-cholera co-infection model is given by the following system of deterministic differential equations (the flow diagram of the model is shown in Figure 1 and the associated variables and parameters of the model are presented in Tables1 and 2, respectively):

**Figure 1:**
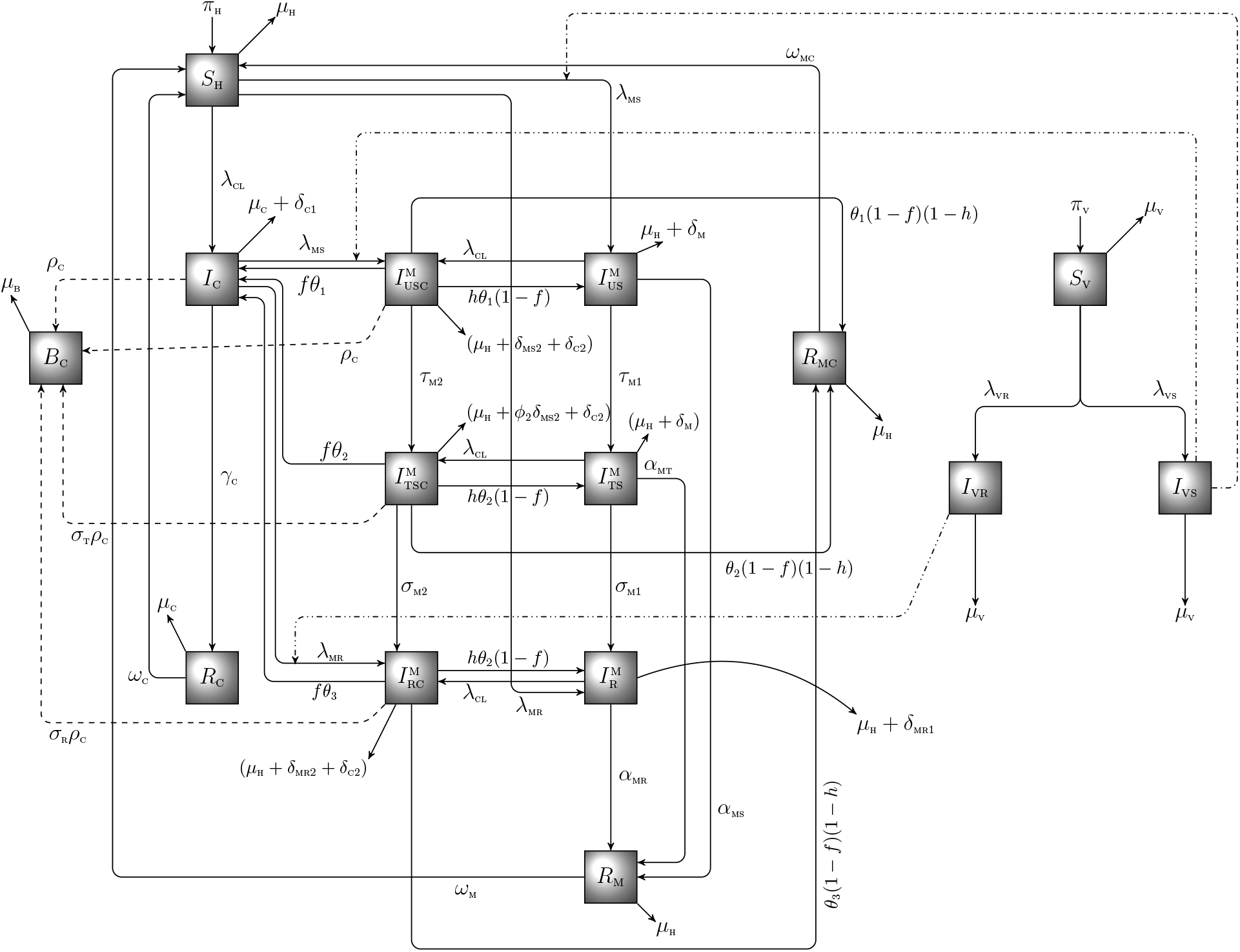
Schematic diagram of the model (2), with 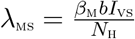, 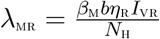, 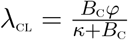, 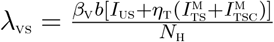, 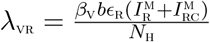

**Table 1:** Description of variables in the model equation

| Variable | Interpretation |
| --- | --- |
| $S_H$ | Susceptible humans |
| $N_H$ | Human population |
| $I_{US}^M$ | Individuals with sensitive malaria strain |
| $I_{TS}^M$ | Individuals treated for malaria infection |
| $R_M$ | Individuals who have recovered from malaria infection |
| $I_C$ | Individuals with cholera infection |
| $R_C$ | Individuals who have recovered from cholera |
| $I_{USC}^M$ | Individuals dually infected with sensitive malaria and Cholera |
| $I_{TSC}^M$ | Individuals who are treated of sensitive malaria and infected with cholera |
| $I_{RC}^M$ | Individuals co-infected with resistant malaria and Cholera |
| $R_{MC}$ | Individuals who have recovered from both malaria and cholera infections |
| $S_V$ | Susceptible vectors |
| $I_{VR}$ | Vectors with resistant malaria strain |
| $I_{VS}$ | Vectors with sensitive malaria strain |
| $B_C$ | Bacteria population |

**Table 2:** Description of parameters in the model (2).

| Parameter | Description | Value (per day) | References |
| --- | --- | --- | --- |
| $\pi_H$ | Recruitment rate for humans | 100 | [24] |
| $\pi_V$ | Recruitment rate for vectors | 1000 | [24] |
| $\beta_M$ | Probability of humans getting infected with malaria | 0.181 | [27] |
| $\beta_V$ | Probability of vectors getting infected with malaria | 0.181 | [27] |
| $b$ | average biting rate for vectors | 0.5 | [26] |
| $\varphi$ | Bacteria contact rate for humans | 0.05 | [13] |
| $\mu_H$ | Natural death rate for humans | $\frac{1}{(70 \times 365)}$ | Assumed |
| $\mu_V$ | Natural death rate for vectors | $\frac{1}{15}, 0.143$ | [24] |
| $\omega_M$ | Malaria waning rate | $\frac{1}{(60 \times 365)}$ | [24] |
| $\omega_C$ | Cholera waning rate | 0.001 | [27] |
| $\omega_{MC}$ | co-infected waning rate | 0.001-0.02 | [13] |
| $\theta_1, \theta_2, \theta_3$ | Recovery rate of co-infected | 0.5 | Assumed |
| $\gamma_C$ | Recovery rate of cholera infected | 0.07 | [27] |
| $\alpha_{MS}$ | Recovery rate from sensitive malaria strain | 0.0078 | [10, 27] |
| $\alpha_{MT}$ | Recovery rate from sensitive malaria strain for treated individuals | 0.1404 | [10, 27] |
| $\alpha_{MR}$ | Recovery rate from resistant malaria strain | 0.0078 | [10, 27] |
| $\eta_T$ | modification parameter for reduced infectiousness of treated individuals | 0.8 | [10] |
| $\eta_R$ | modification parameter for reduced infectiousness of malaria resistant individuals | variable | |
| $\tau_{M1}, \tau_{M2}$ | Rate of administration of antimalarial drugs | variable | |
| $\sigma_{M1}, \sigma_{M2}$ | rate of resistance development to antimalarial drugs | variable | |
| $f$ | fraction of co-infected who recover from malaria only | 0.2 | Assumed |
| $h$ | fraction of co-infected who recover from cholera only | 0.5 | Assumed |
| $\rho_C$ | cholera infected contribution to aquatic | 0.7 | Assumed |
| $\sigma_T, \sigma_R$ | modification parameters | 0.9 | Assumed |
| $\delta_{MS1}, \delta_{MS2}$ | Malaria induced death rates | 0.05-0.1 | [13, 25] |
| $\delta_{MT}$ | Malaria induced death rates for treated individuals | 0.05 | Assumed |
| $\delta_{C1}, \delta_{C2}$ | Cholera induced death rates | 0.0001 | Assumed |
| $\phi_1, \phi_2$ | Modification parameters | 0.7 | Assumed |

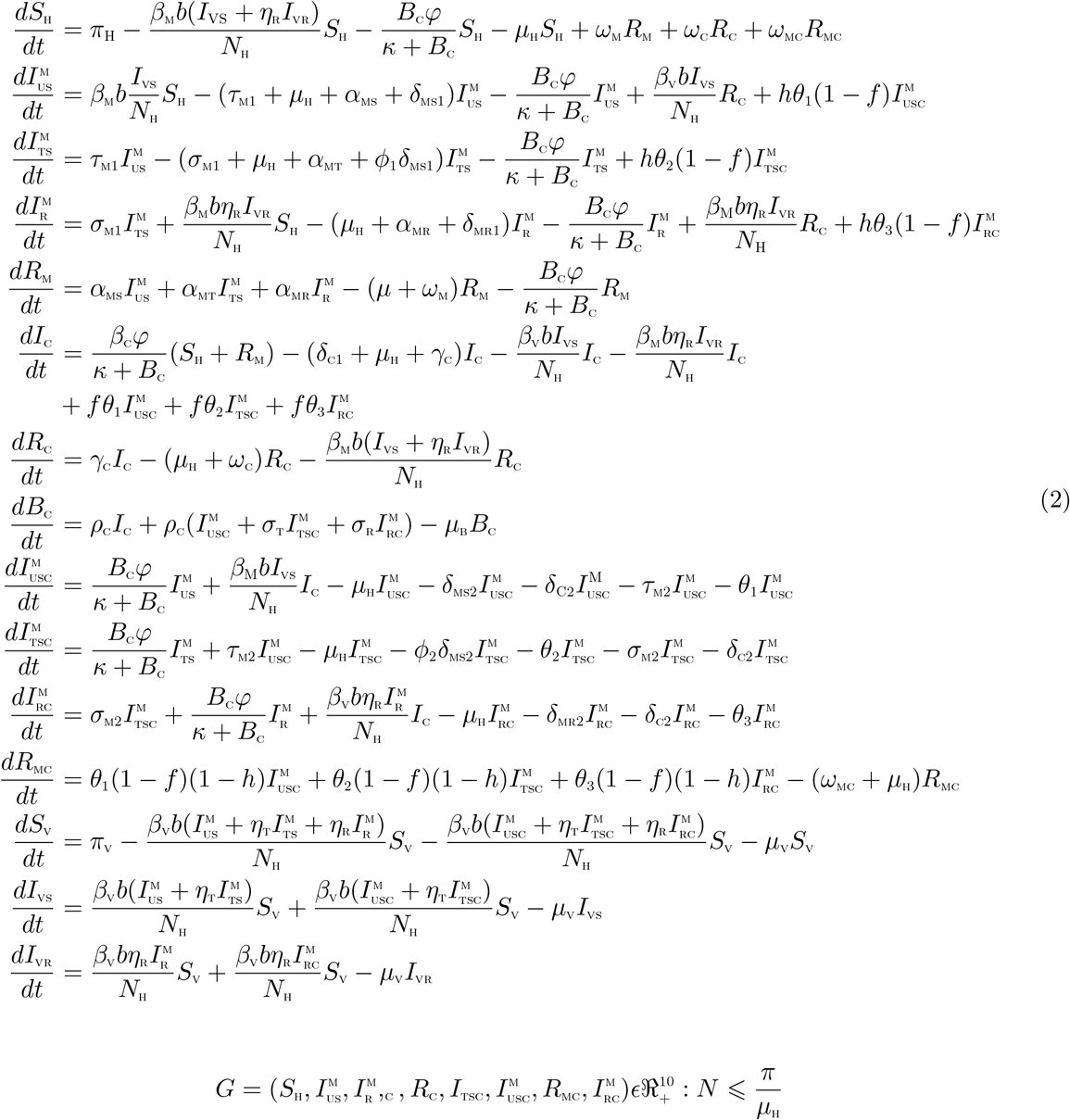

## 3 Mathematical analysis of the co-infection model

In this section, we qualitatively analyze the model (2) to better understand the dynamics of the co-infection of drug-resistant Malaria and cholera in a population.

### 3.1 Basic properties of the Malaria-Cholera co-infection model

### 3.2 Positivity and boundedness of solutions

For the model (2) to be epidemiologically meaningful, it is important to prove that all its state variables are non-negative for all time (*t*). In other words, solutions of the model system (2) with positive initial data will remain positive for all time t > 0.

#### Theorem 3.1

*Let the initial data S*_H_ > 0, 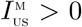, 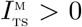, 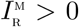, *R*_M_ > 0, *I*_C_ > 0, *R*_C_ > 0, *B*_C_ > 0, 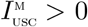, 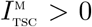, 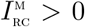, *R*_MC_ > 0, *S*_V_ > 0, *I*_VS_ > 0, *I*_VR_ > 0. *Then the solution of the model are positive for all t* > 0

##### Proof

Let

*t*_1_ = sup{*t* > 0: *S*_H_ > 0, 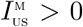, 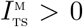, 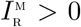, 0, *R*_M_ > 0, *I*C > 0;*R*C > 0;*B*C > 0; 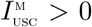, 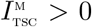, 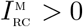, *R*_MC_ > 0, *S*V > 0; *I*VS > 0; *I*VR > 0 ∈ [0; *t*]}. Thus, *t*_1_ > 0.

We have, from the first equation of the system (2) that

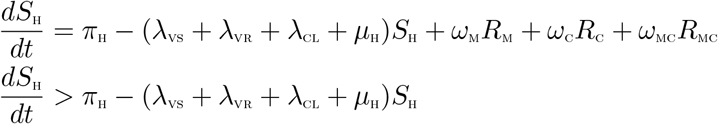

which can be re-written as

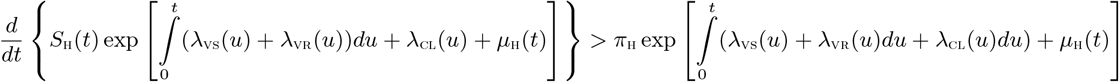

Hence:

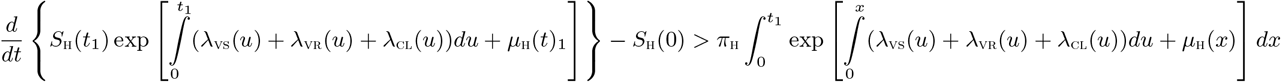

so that

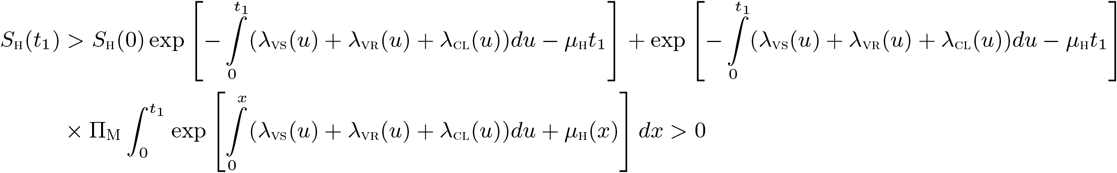

Similarly, it can be shown that:

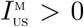, 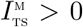, 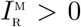, *R*_M_ > 0, *I*_C_ > 0, *R*_C_ > 0, *B*_C_ > 0, 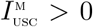, 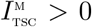, 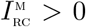, *R*_MC_ > 0, *S*_V_ > 0, *I*_VS_ > 0, *I*_VR_ > 0.

### 3.3 Invariant regions

The co-infection model (2) will be analyzed in a biologically feasible region as follows. We first show that the system (4) is dissipative in a proper subset 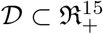. The system (2) is split into two parts, namely the human population (*N*_H_) (with 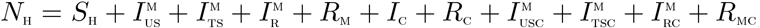), the bacteria population, *B*_C_ and the vector population, (*N*_V_) (with *N*_V_ = *S*_V_ + *I*_VS_ + *I*_VR_).

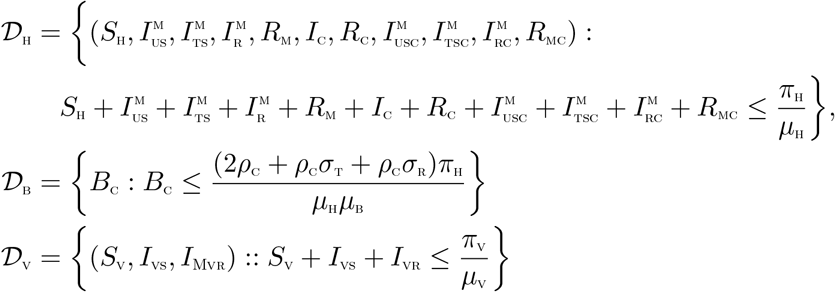

The following steps are followed to establish the positive invariance of 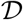 (i.e. solutions in 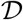 remain in 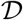 for all time *t* > 0).

Adding human, bacteria and vector components of the differential system (2), respectively, gives

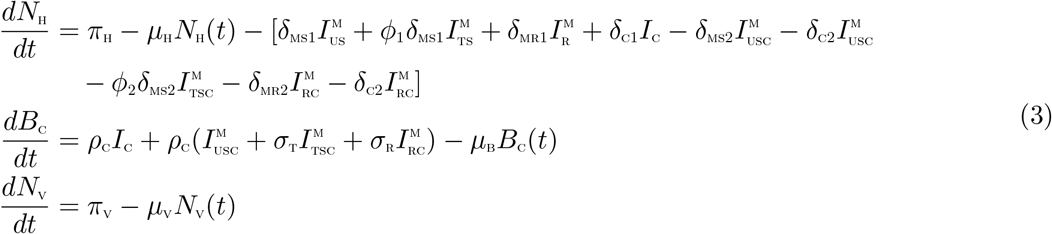

From (3), we have that

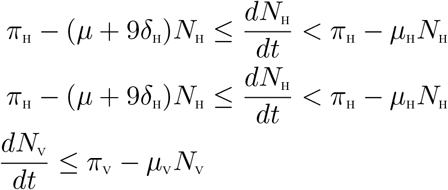

where *δ*_H_ = min{*δ*_MS1_*, ϕ*_1_*δ*_MS1_, *δ*_MR1_*, δ*_C1_, *δ*_MS2_, *δ*_C2_, *ϕ*_2_*δ*_MS2_, *δ*_MR2_}

Using the Comparison theorem [19], we have that,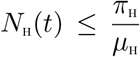 and 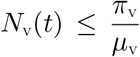 if 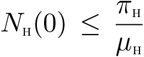 and 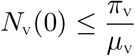 respectively. Thus, the region 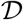 is positively invariant. Hence, it is sufficient to consider the dynamics of the flow generated by the system (2) in 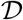. In this region, the model can be considered as being epidemiologically and mathematically well-posed [20]. Thus, every solution of the model (2) with initial conditions in 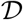 remains in 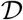 for all time *t* ≥ 0. Therefore, the *ω*-limit sets of the system (2) are contained in 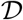. Thus result is summarized thus.

#### Lemma 3.1

*The region* 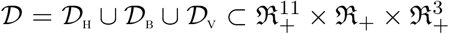 *is positively-invariant for the model* (2) *with initial conditions in* 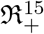.

### 3.4 Basic reproduction number of the co-infection model (2)

The Malaria-Cholera co-infection model (2) has a DFE, obtained by setting the right-hand sides of the equations in the model (2) to zero, given by

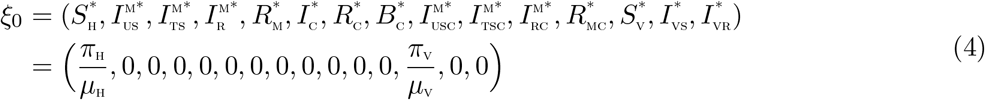

The matrices *F* and *V*, for the new infection terms and the remaining transfer terms, evaluated at the disease free equilibrium (DFE) are, respectively, given by

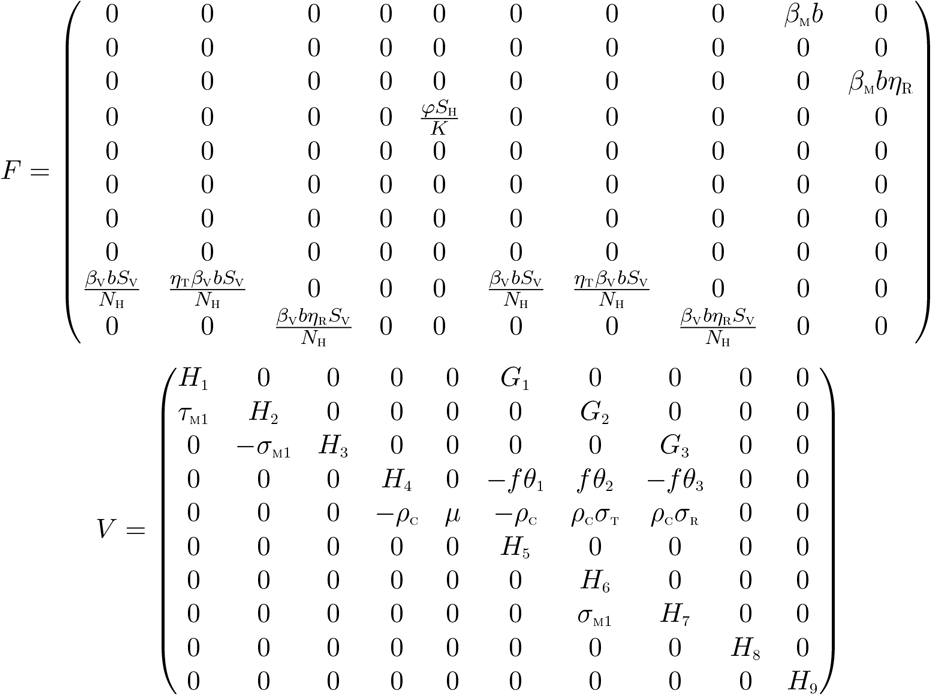

where

*H*_1_ = *τ*_M1_ + *µ*_H_ + *α*_MS_ + *δ*_MS1_, *H*_2_ = *σ*_M1_ + *µ*_H_ + *α*_MT_ + *ϕ*_1_*δ*_MS1_, *H*_3_ = *µ*_H_ + *α*_MR_ + *δ*_MR1_, *H*_4_ = *δ*_C1_ + *µ*_H_ + γ_C_ *H*_5_ = *µ*_H_ + *δ*_MS2_ + *δ*_C2_ + *τ*_M2_ + *θ*_1_, *H*_6_ = *ϕ*_1_*δ*_MS2_ + *µ*_H_ + *θ*_2_*σ*_M2_ + *δ*_C2_, *H*_7_ = *δ*_MR2_ + *µ*_H_ + *δ*_C2_ + *θ*_3_

The basic reproduction number of the Malaria-Cholera co-infection model (2), using the approach illustrated in van den Driessche and Watmough [21], is given by 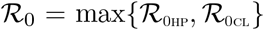 where 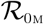 and 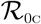 are, respectively, the Malaria and Cholera associated reproduction numbers, given by

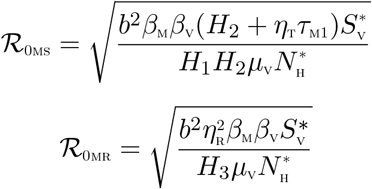

and

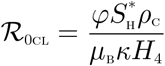

### 3.5 Local asymptotic stability of disease-free equilibrium (DFE) of the co-infection model (2)

#### Lemma 3.2

*The DFE*, ξ_0_, *of the Malaria-Cholera co-infection model* (2) *is locally asymptotically stable if* 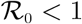, and *unstable if* 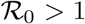.

##### Proof

The local stability of the co-infection model is analysed by the Jacobian matrix of the system (2) at ξ_0_, given by:

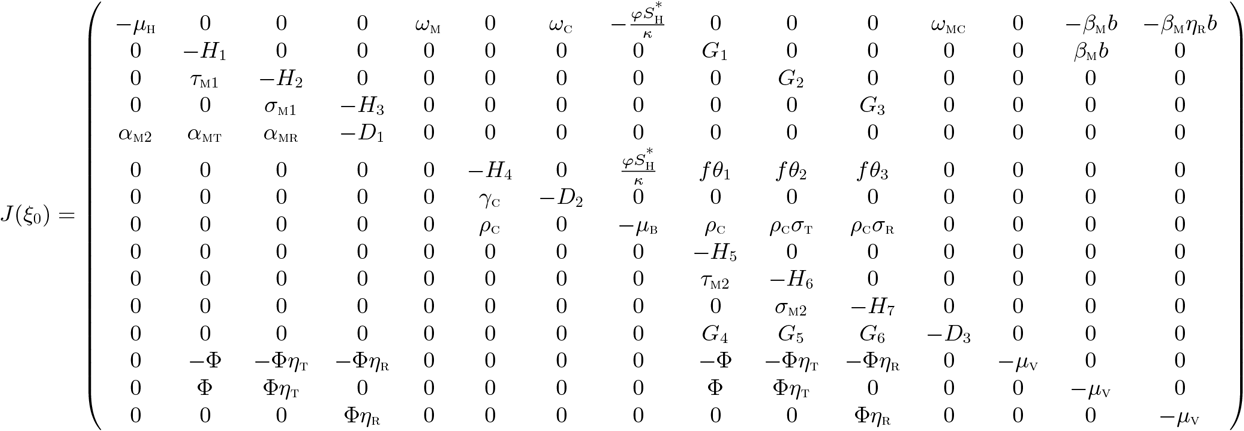

with,

*H*_1_ = *τ*_M1_ + *µ*_H_ + *α*_MS_ + *δ*_MS1_, *H*_2_ = *σ*_M1_ + *µ*_H_ + *α*_MT_ + *ϕ*_1_*δ*_MS1_, *H*_3_ = *µ*_H_ + *α*_MR_ + *δ*_MR1_, *H*_4_ = *δ*_C1_ + *µ*_H_ + γ_C_ *H*_5_ = *µ*_H_ + *δ*_MS2_ + *δ*_C2_ + *τ*_M2_ + *θ*_1_, *H*_6_ = *ϕ*_1_*δ*_MS2_ + *µ*_H_ + *θ*_2_*σ*_M2_ + *δ*_C2_, *H*_7_ = *δ*_MR2_ + *µ*_H_ + *δ*_C2_ + *θ*_3_

*G*_1_ = *hθ*_1_(1 − *f*), *G*_2_ = *hθ*_2_(1 − *f*),*G*_3_ = *hθ*_3_(1 − *f*), *G*_4_ = *θ*_1_(1 − *f*)(1 − *h*), *G*_5_ = *θ*_2_(1 − *f*)(1 − *h*) *G*_6_ = *θ*_3_(1 − *f*)(1 − *h*), *D*_1_ = *µ*_H_ + *ω*_M_, *D*_2_ = *µ*_H_ + *ω*_C_, *D*_3_ = *µ*_H_ + *ω*_MC_, 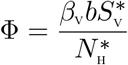

The eigenvalues are λ_1_ = −*D*_1_, λ_1_ = −*D*_2_, λ_3_ = −*D*_3_, λ_4_ = −*H*_5_, λ_5_ = −*H*_6_, λ_6_ = −*H*_7_, λ_7_ = −*µ*_H_, λ_8_ = −*µ*_V_ and the solutions of the characteristic polynomials:

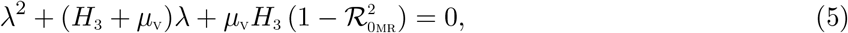

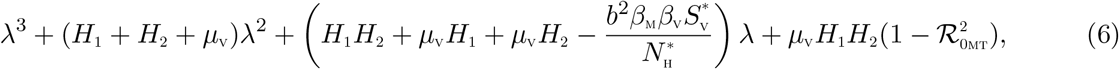

and

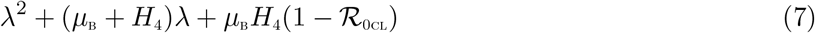

Applying the Routh-Hurwitz criterion, the quadratic equations (5), (6) and (7) will have roots with negative real parts if and only if 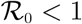. As a result, the disease-free equilibrium, ξ_0_ is locally asymptotically stable if 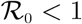.

### 3.6 Global asymptotic stability(GAS) of the disease-free equilibrium(DFE) ξ_0_ of the co-infection model (2)

The approach illustrated in [22] is used to investigate the global asymptotic stability of the disease free equilibrium of the co-infection model. In this section, we list two conditions that if met, also guarantee the global asymptotic stability of the disease-free state. First, system (2) must be written in the form:

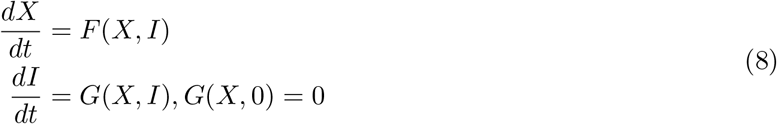

where *X* ∈ *R^m^* denotes (its components) the number of uninfected individuals and *I* ∈ *R^n^* denotes (its components) the number of infected individuals including latent, infectious, etc. *U*_0_ = (*X*^*^, 0) denotes the disease-free equilibrium of this system. The conditions (*H*1) and (*H*2) below must be met to guarantee local asymptotic stability:

(H1): For 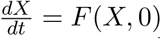, *X*^*^is globally asymptotically stable (GAS), (H2): *G*(*X, I*) = *AI* − Ĝ(*X, I*)*X*, *G*(*X*, *I*) ≥ 0 for (*X*, *I*) ∈ Ω,

where *A* = *D*_1_*G*(*X*^*^, 0) is an M-matrix (the off-diagonal elements of A are nonnegative) and Ω is the region where the model makes biological sense. If System (2) satisfies the above two conditions then the following theorem holds:

#### Theorem 3.2

*The fixed point U*_0_ = (*X*^*^, 0) *is a globally asymptotic stable (GAS) equilibrium of* (2) *provided that R*_0_ < 1 *(LAS) and that assumptions* (*H*1) *and* (*H*2) *are satisfied*

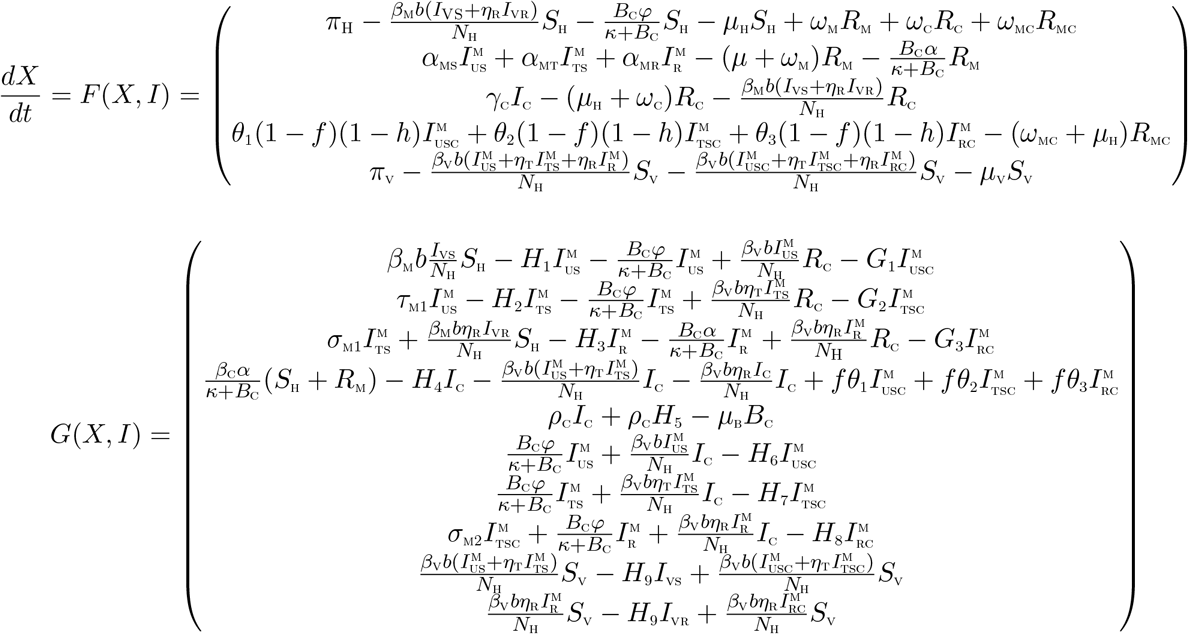

where X denotes the number of non-infectious individuals and *I* denotes the number of infected individuals.

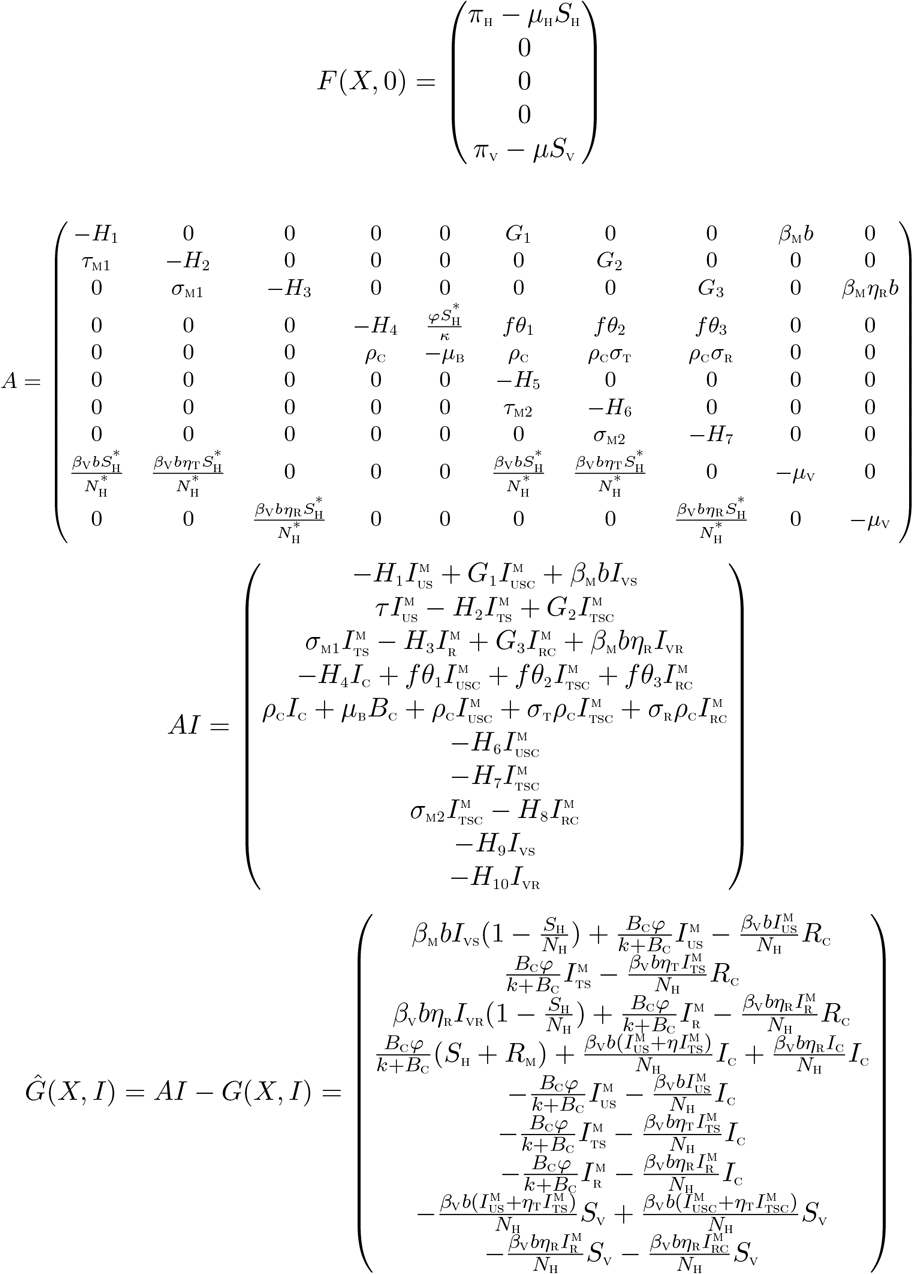

It is clear from the above, that, *Ĝ*(*X, I*) ≱ 0. Hence the DFE may not be globally asymptotically stable, suggesting the possibility of a backward bifurcation.

### 3.7 Backward bifurcation analysis of the full co-infection model (2)

In this section, we shall seek to determine the type of bifurcation the model (2) will exhibit, using the approach illustrated by Castillo-Chavez and Song [23]. We establish the result below

#### Theorem 3.3

*Suppose a backward bifurcation coefficient a* > 0, *(with a defined below), when* 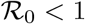

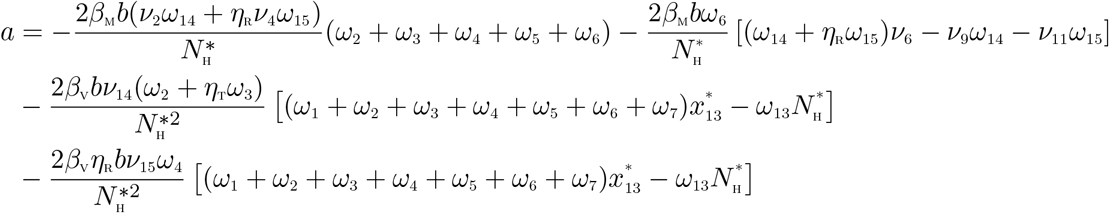

*then model* (2) *undergoes the phenomenon of backward bifurcation at* 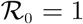. *If a* < 0, then *the system* (2) *exhibits a forward bifurcation at* 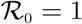.

##### Proof

Suppose

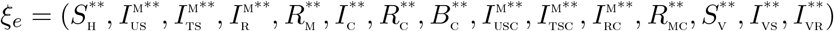

represents any arbitrary endemic equilibrium of the model. The existence of backward bifurcation will be studied using the Centre Manifold Theory by Castillo-Chavez and Song [23]. To apply this theory, it is appropriate to do the following change of variables.

Let

*S*_H_ = *x*_1_, 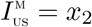, 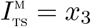, 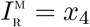, *R*_M_ = *x*_5_, *I*_C_ = *x*_6_, *R*_C_ = *x*_7_, *B*_C_ = *x*_8_, 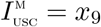, 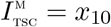, 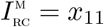, *R*_MC_ = *x*_12_, *S*_V =_ *x*_13_, *I*_VS =_ *x*_14_, *I*_VR =_ *x*_15_

Moreover, using the vector notation

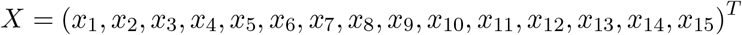

the model (2) can be re-written in the form

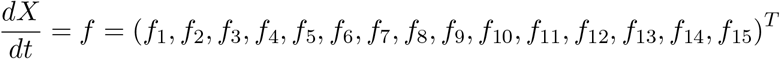

as follows:

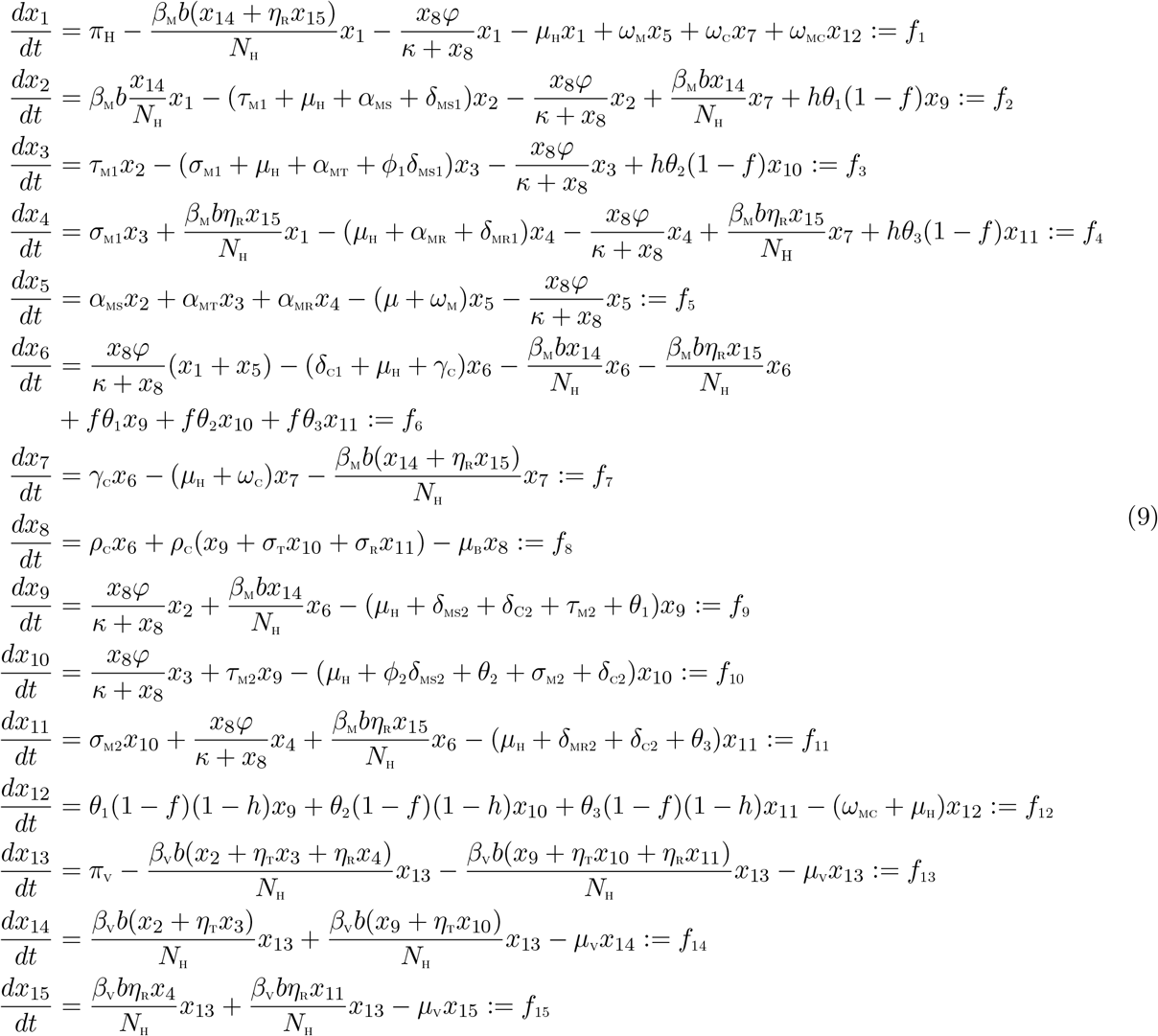

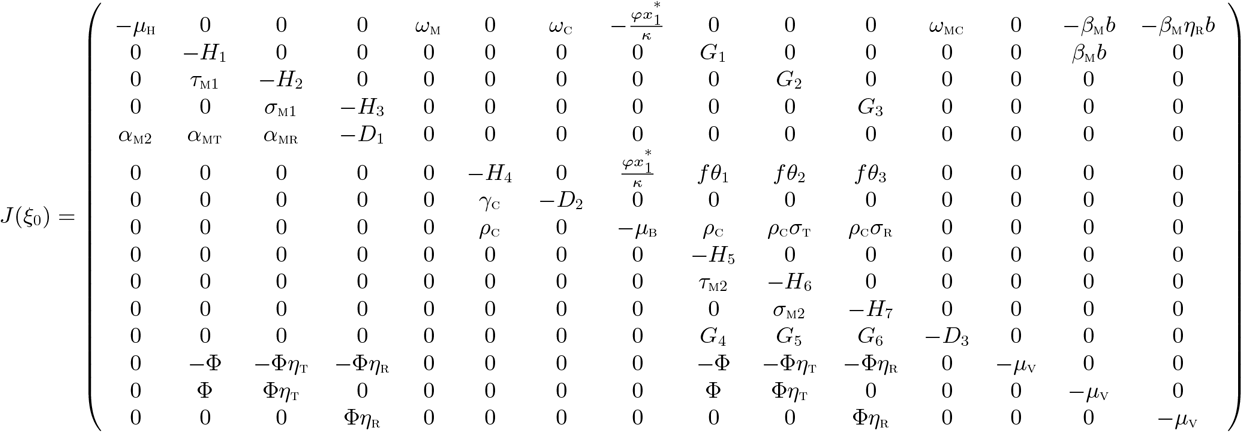

with,

*H*_1_ = *τ*_M1_ + *µ*_H_ + *α*_MS_ + *δ*_MS1_, *H*_2_ = *σ*_M1_ + *µ*_H_ + *α*_MT_ + *ϕ*_1_*δ*_MS1_, *H*_3_ = *µ*_H_ + *α*_MR_ + *δ*_MR1_, *H*_4_ = *δ*_C1_ + *µ*_H_ + γ_C_ *H*_5_ = *µ*_H_ + *δ*_MS2_ + *δ*_C2_ + *τ*_M2_ + *θ*_1_, *H*_6_ = *ϕ*_1_*δ*_MS2_ + *µ*_H_ + *θ*_2_*σ*_M2_ + *δ*_C2_, *H*_7_ = *δ*_MR2_ + *µ*_H_ + *δ*_C2_ + *θ*_3_

*G*_1_ = *hθ*_1_(1 − *f*), *G*_2_ = *hθ*_2_(1 − *f*),*G*_3_ = *hθ*_3_(1 − *f*), *G*_4_ = *θ*_1_(1 − *f*)(1 − *h*), *G*_5_ = *θ*_2_(1 − *f*)(1 − *h*) *G*_6_ = *θ*_3_(1 − *f*)(1 − *h*), *D*_1_ = *µ*_H_ + *ω*_M_, *D*_2_ = *µ*_H_ + *ω*_C_, *D*_3_ = *µ*_H_ + *ω*_MC_, 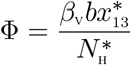

Consider the case when 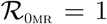. Assume, further, that thee product *β*_M_*η*_R_ is chosen as a bifurcation parameter. Solving for *β*_M_*η*_R_ = *β*^*^ from 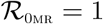 gives

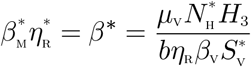

Evaluating the Jacobian of the system (9) at the DFE, *J*(ξ_0_), and using the approach in Castillo-Chavez and Song [23], we have that *J*(ξ_0_) has a right eigenvector (associated with the non-zero eigenvalue) given by

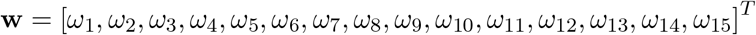

where,

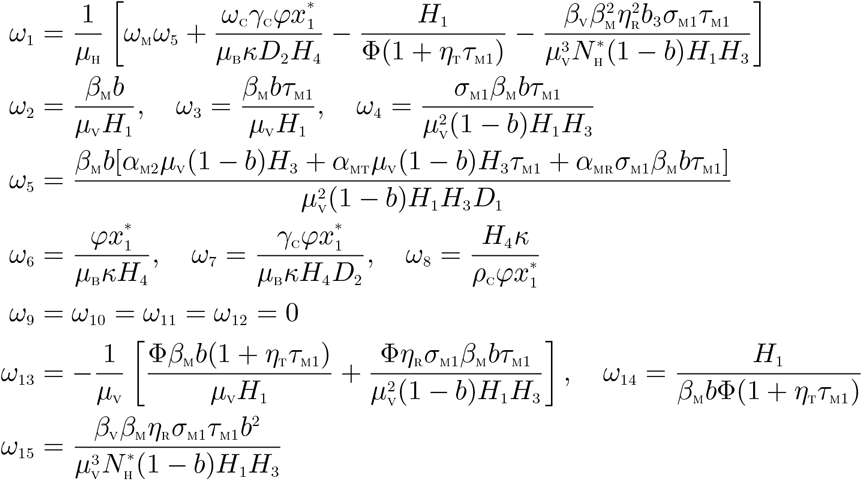

Similarly, the components of the left eigenvector of 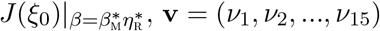, satisfying v.w = 1, are given by:

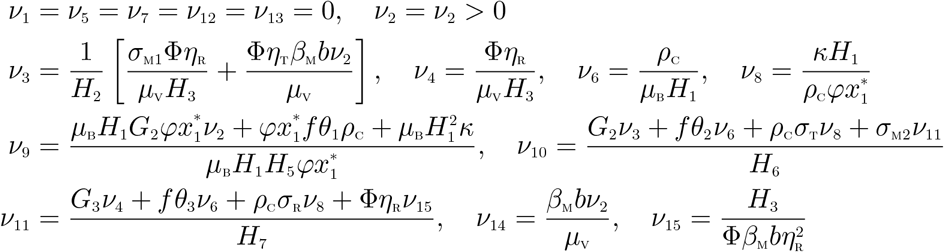

The non-zero second partial derivatives of the functions *f_i_*(*i* = 1,…, 15) are given by

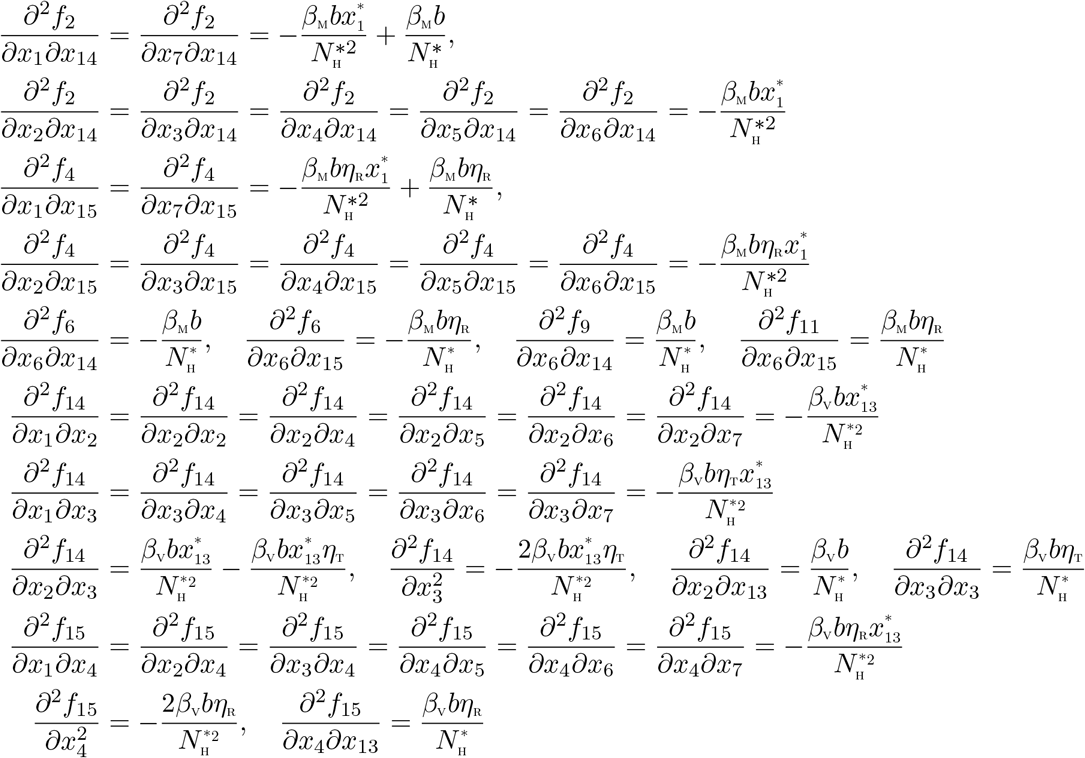

The associated bifurcation coefficients defined by *a* and *b*, given by:

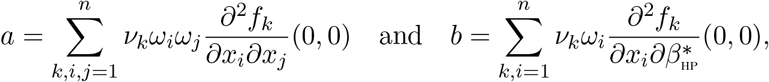

are computed to be

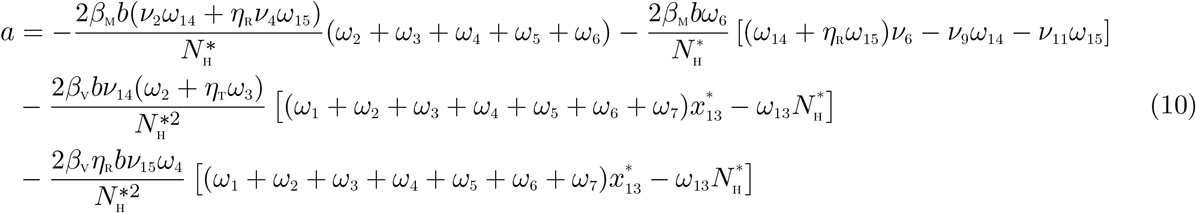

and

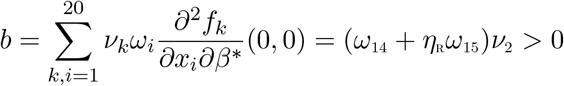

Since the bifurcation coefficient b is positive, it follows from Theorem 4.1 in Castillo-Chavez and Song [23] that the model (2), or the transformed model (9), will undergo the phenomenon of backward bifurcation if the coefficient, a, given by (10) is positive.

## 4 Analysis of the optimal control model

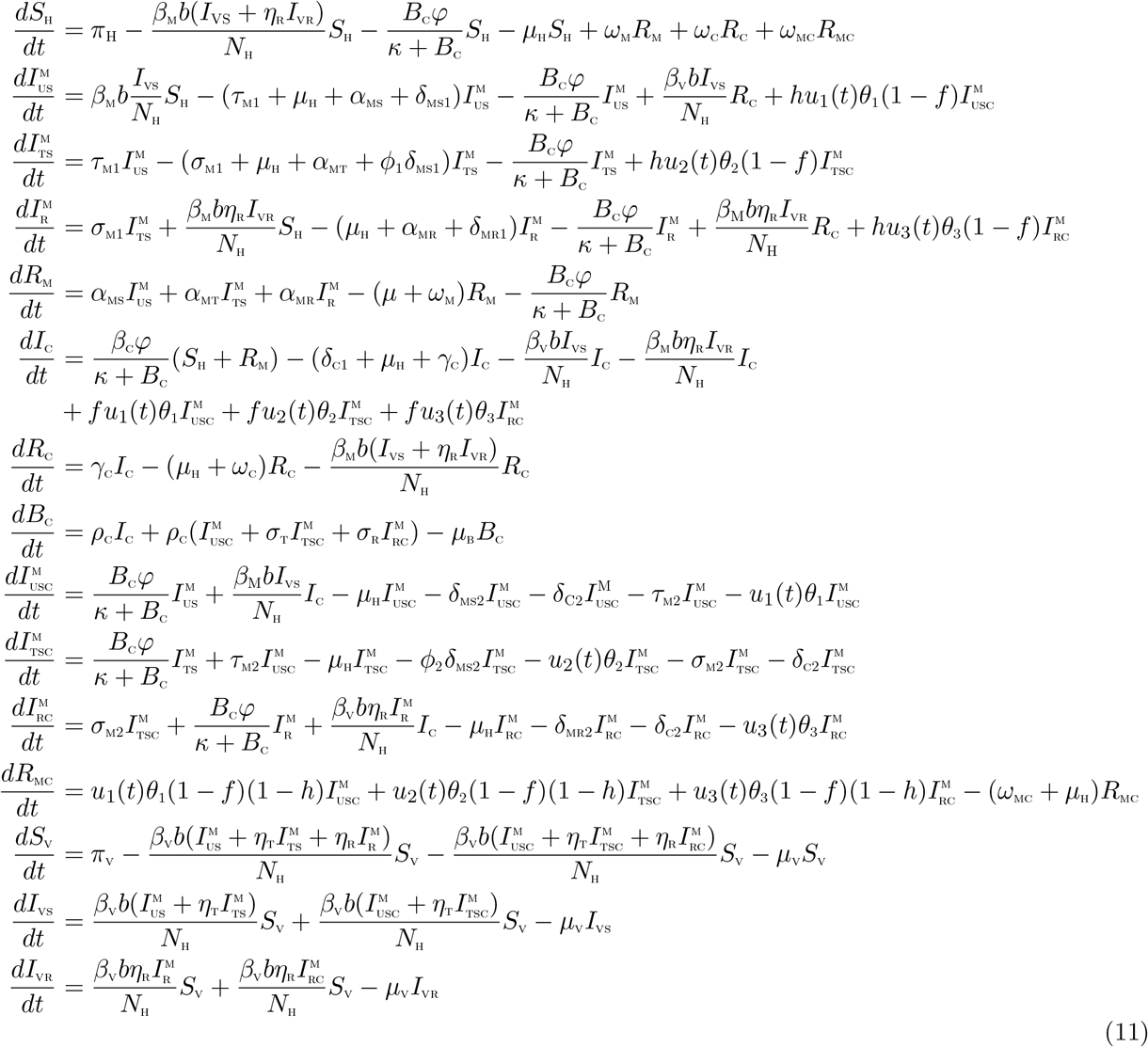

The control functions, *u*_1_(*t*), *u*_2_(*t*), and *u*_3_(*t*) are bounded, Lebesgue integrable functions. The control *u*_1_(*t*) represents treatment efforts for co-infected individuals in 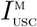 compartment. *u*_2_(*t*) represents treatment efforts for co-infected individuals in 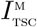 compartment. The control *u*_3_(*t*) represents treatment efforts for co-infected individuals in 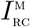 compartment. The controls *u*_1_, *u*_2_,*u*_3_ satisfies 0 ≤ *u*_1_,*u*_2_,*u*_3_ ≤ 1. The optimal control system examines scenarios where the number of co-infected cases and the cost of implementing the controls *u*_1_(*t*),*u*_2_(*t*), and *u*_3_(*t*) are minimized subject to the state system (11). For this, we consider the objective functional

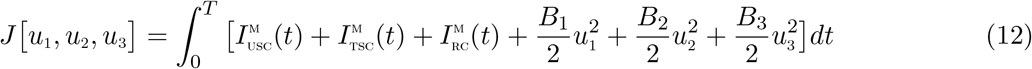

*T* is the final time. We seek to find an optimal control, 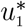, 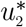, 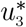, such that

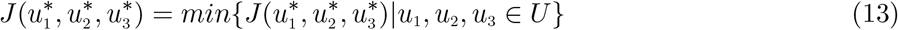

where 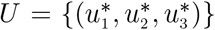 such that 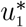, 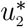, 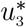 are measurable with 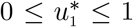, 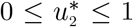, 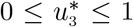, for *t* ∈ [0, *T*] is the control set. The Pontryagin’s Maximum Principle [28] gives the necessary conditions which an optimal control pair must satisfy. This principle transforms (11), (12) and (13) into a problem of minimizing a Hamiltonian, 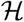, pointwisely with regards to the control functions, *u*_1,_ *u*_2_, *u*_3_:

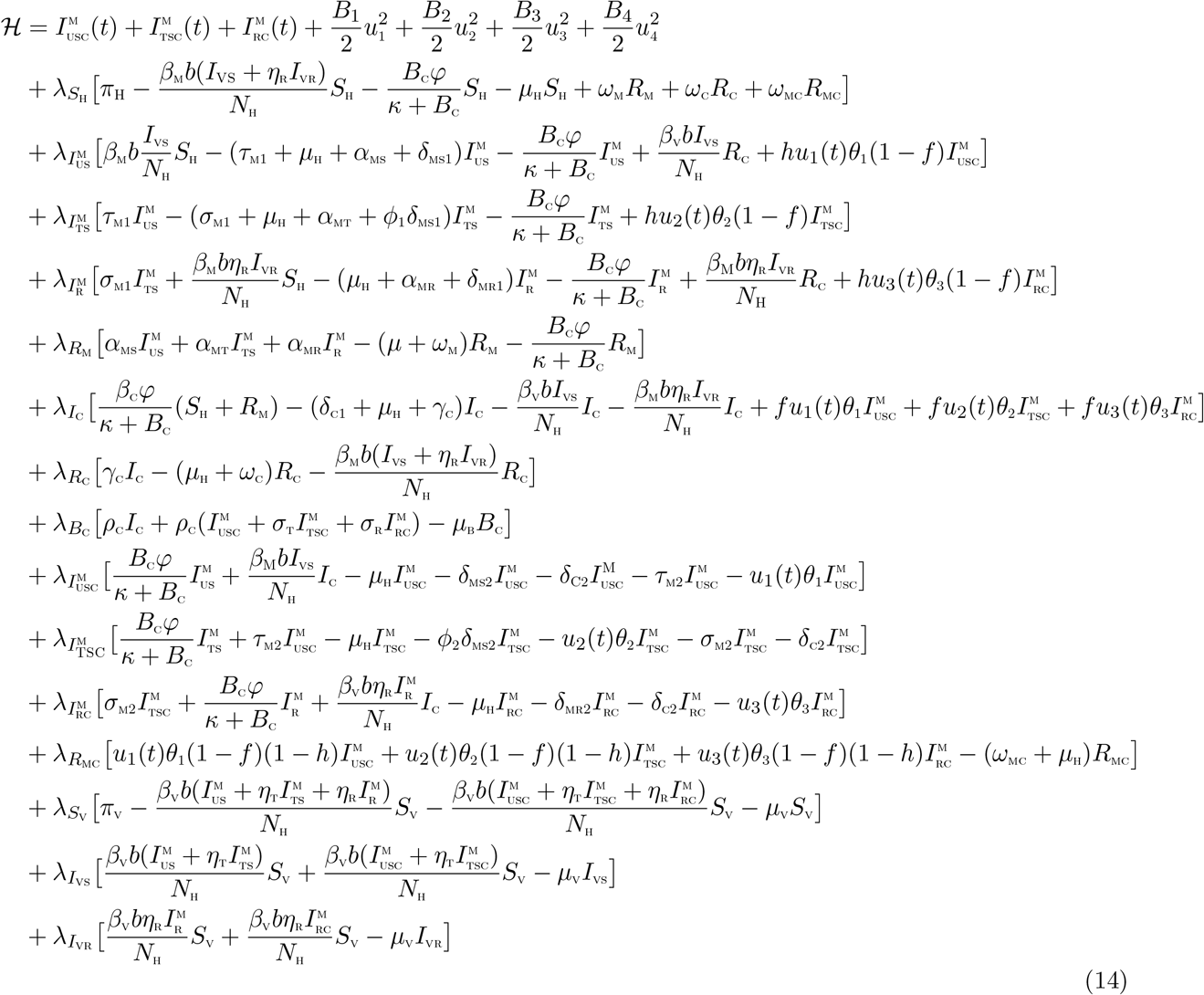

### Theorem 4.1

*For an optimal control set u*_1;_ *u*_2_, *u*_3_ *that minimizes J over U, there are adjoint variables*, λ_1;_ λ_2_,…, λ_15_ *satisfying*

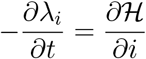

*and with transversality conditions*

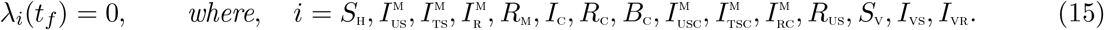

*Furthermore*,

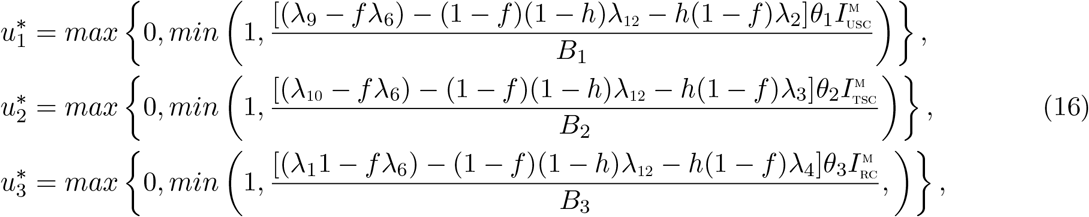

#### Proof of Theorem 4.1

Suppose 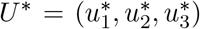 is an optimal control and *S*_H_, 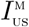, 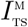, 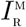, *R*_M_, *I*_C_, *B*_C_, 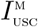, 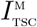, 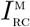, *R*_US_, *S*_V_, *I*_VS_, *I*_VR_. are the corresponding state solutions. Applying the Pontryagin’s Maximum Principle [28], there exist adjoint variables satisfying:

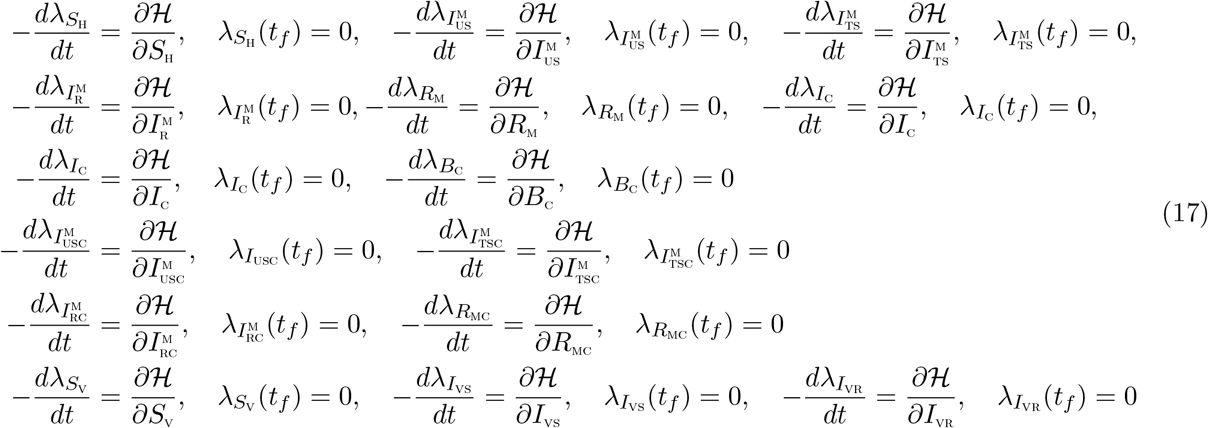

with transversality conditions;

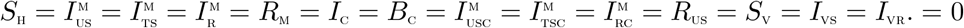 We can determine the behaviour of the control by differentiating the Hamiltonian, 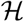 with respect to the controls(*u*_1_, *u*_2_, *u*_3_, *u*_4_) at *t*. On the interior of the control set, where 0 < *u_j_* < 1 for all (*j* = 1,2, 3, 4), we obtain

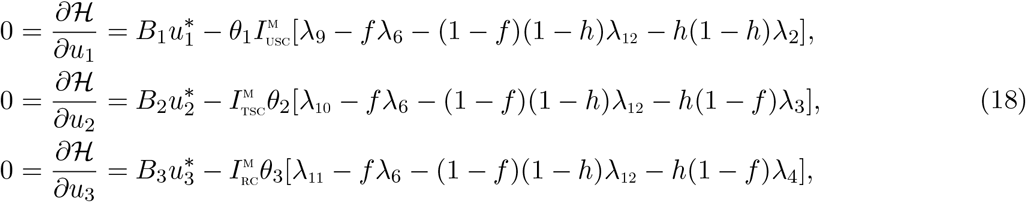

Therefore, we have that [29]

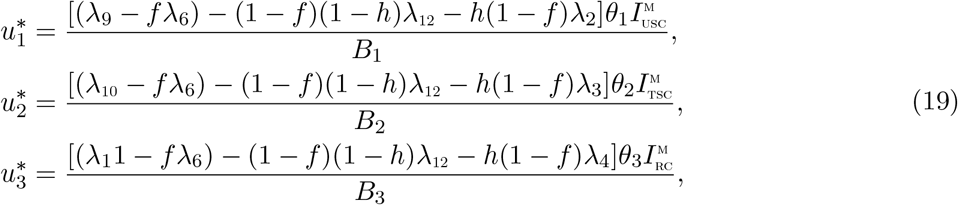

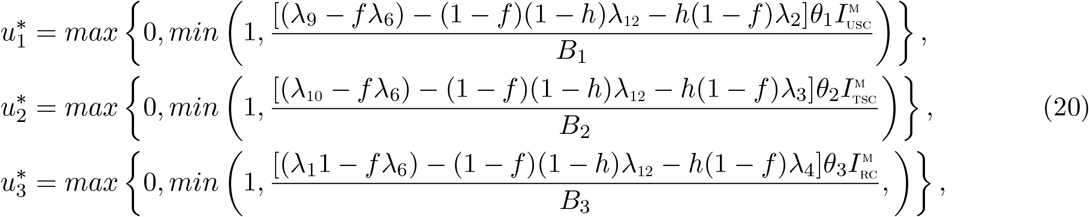

## 5 Numerical simulations

Figure 2 shows the simulation of the total number of individuals untreated of sensitive strain and coinfected with cholera, when there is no drug resistance. it is observed that a total of 1,590 co-infected cases were averted. However as presented in Figure 3 when there is drug resistance, a total of 237 co-infection cases were averted. figure 4 shows the simulations of the total number of individuals treated of sensitive malaria strain and co-infected with cholera. It is observed that a total of 2,527 co-infection cases were prevented, when there is no resistance. However, when there is malaria drug resistance, as depicted figure 5, a total of 116 co-infection cases were averted. Figure 6 depicts the simulations of the total number of individuals co-infected with resistant malaria and cholera. It is seen that in the absence of malaria drug resistance, the treatment control applied prevented a total of 36,350 co-infection cases. However, in the presence of malaria drug resistance, as shown Figure 7 a total of 40,590 co-infection cases were averted when the treatment control is applied. It is imperative to note that in the presence of treatment controls, more co-infection cases were averted when there is drug resistance, than whne there is no drug resustance, for co-infected individuals in compartments 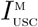 and 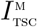. However, it is observed that for co-infected individuals in compartment 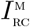, more co-infection cases were averted in the presence of malaria drug resistance than when there is no drug resistance, when treatment controls are applied in both cases. This points to the fact that malaria drug resistance can greatly influence the co-infection cases averted, even in the presence of treatment controls for co-infected individuals.

**Figure 2:**
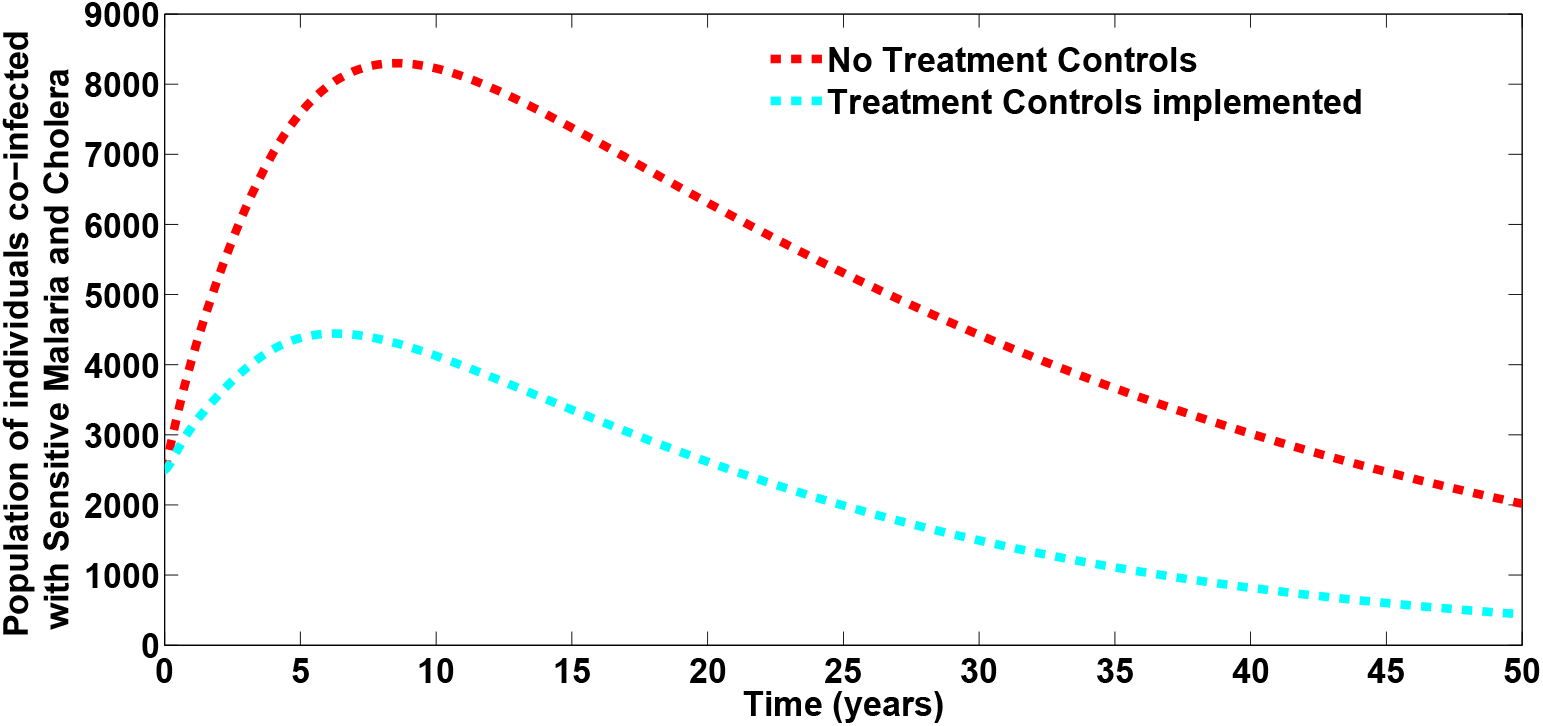
Simulations of the model (2) showing the total number of untreated of sensitive strain and co-infected with cholera, when there is no drug resistance. Here, *σ*_M1_ = *σ*_M2_ = 0. All other parameters as in Table 2

**Figure 3:**
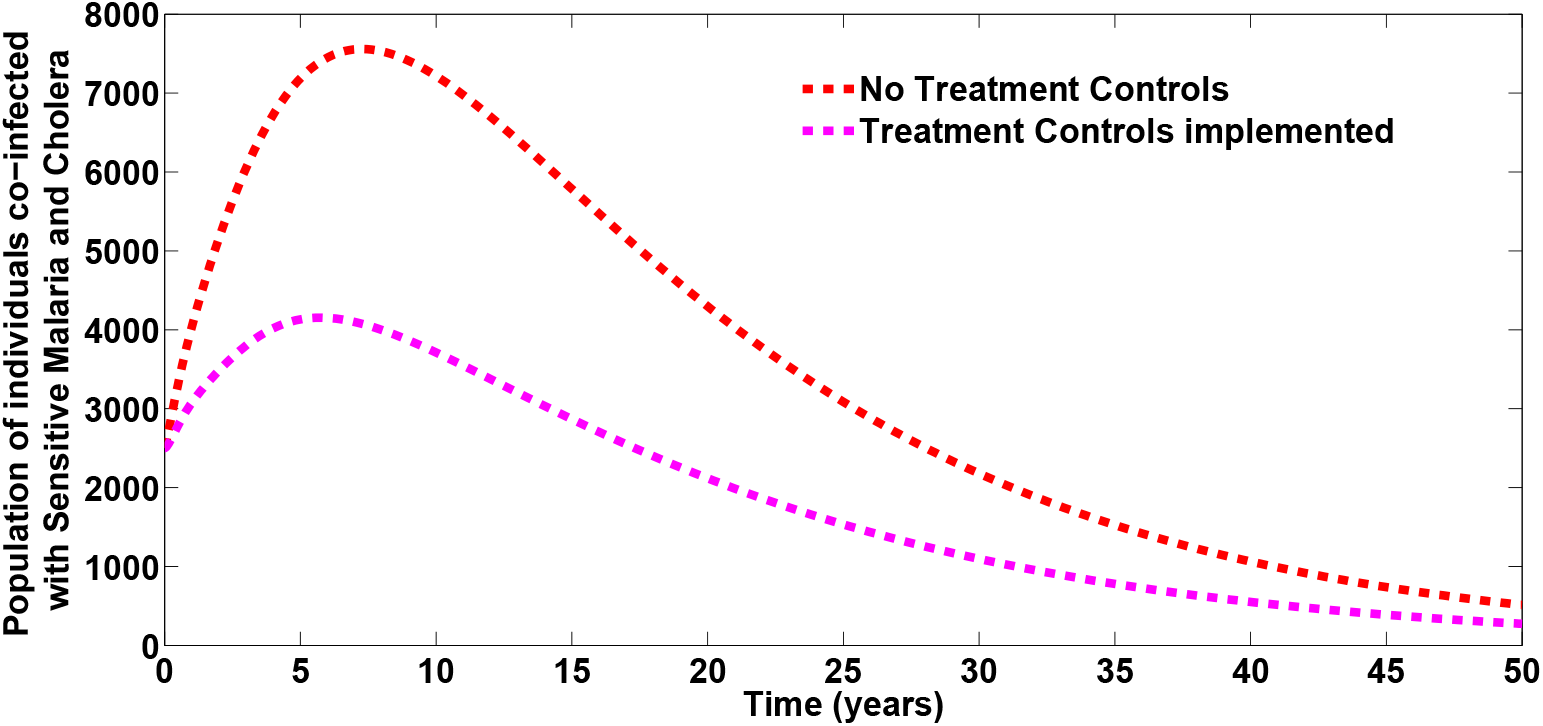
Simulations of the model (2) showing the total number of untreated of sensitive strain and co-infected with cholera, when there is drug resistance. Here, *σ*_M1_ = *σ*_M2_ = 0.4. All other parameters as in Table 2

**Figure 4:**
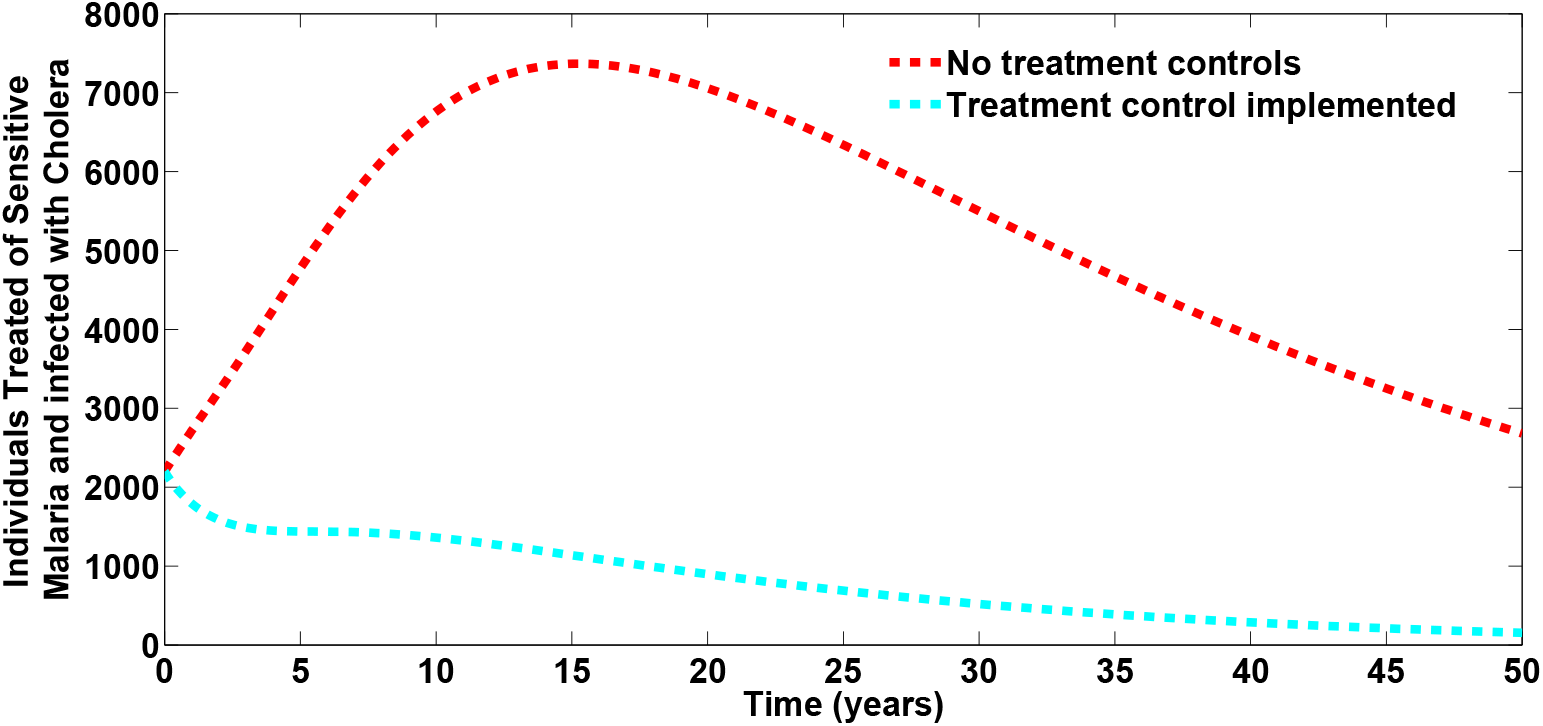
Simulations of the model (2) showing the total number of treated of sensitive strain and coinfected with cholera, when there is no drug resistance. Here, *σ*_M1_ = *σ*_M2_ = 0. All other parameters as in Table 2

**Figure 5:**
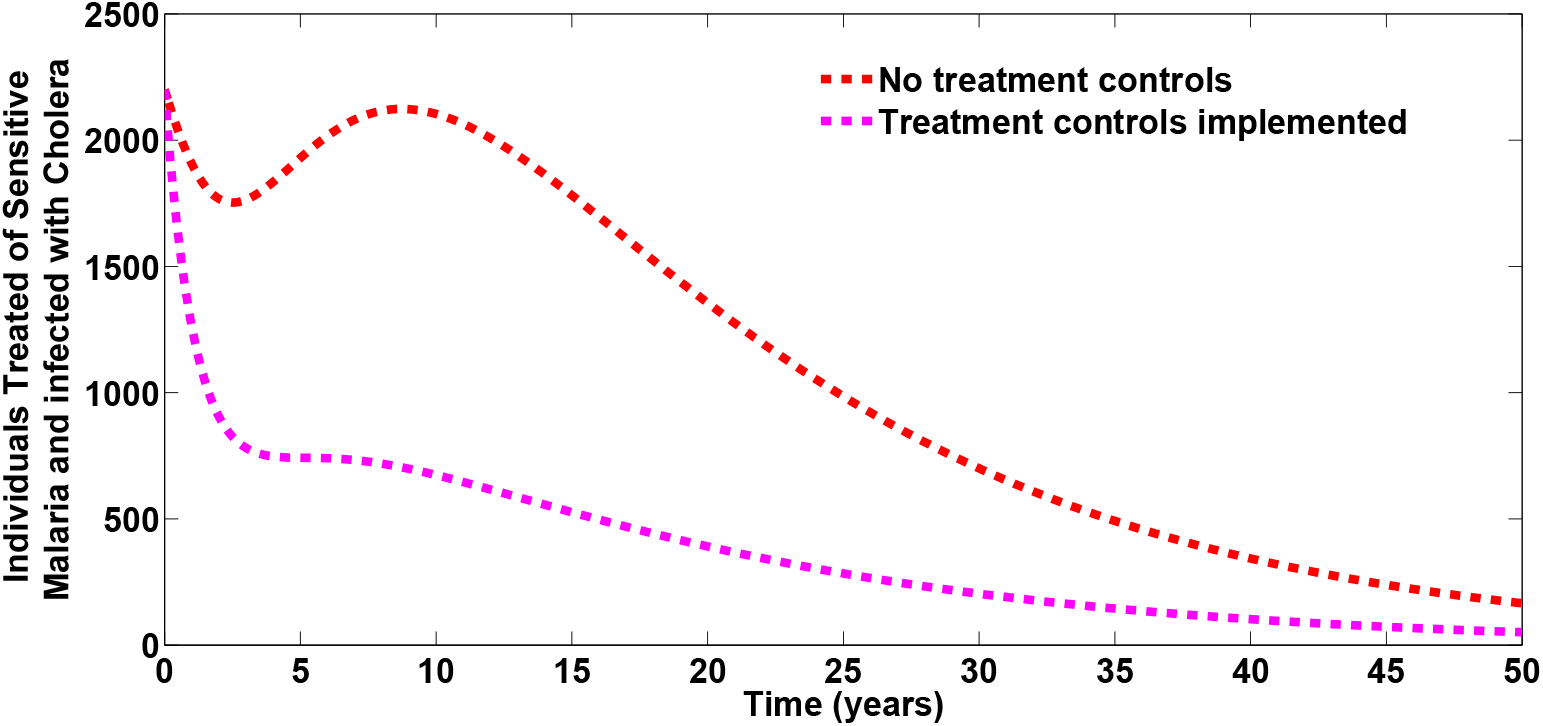
Simulations of the model (2) showing the total number of treated of sensitive strain and co-infected with cholera, when there is drug resistance. Here, *σ*_M1_ = *σ*_M2_ = 0.4. All other parameters as in Table 2

**Figure 6:**
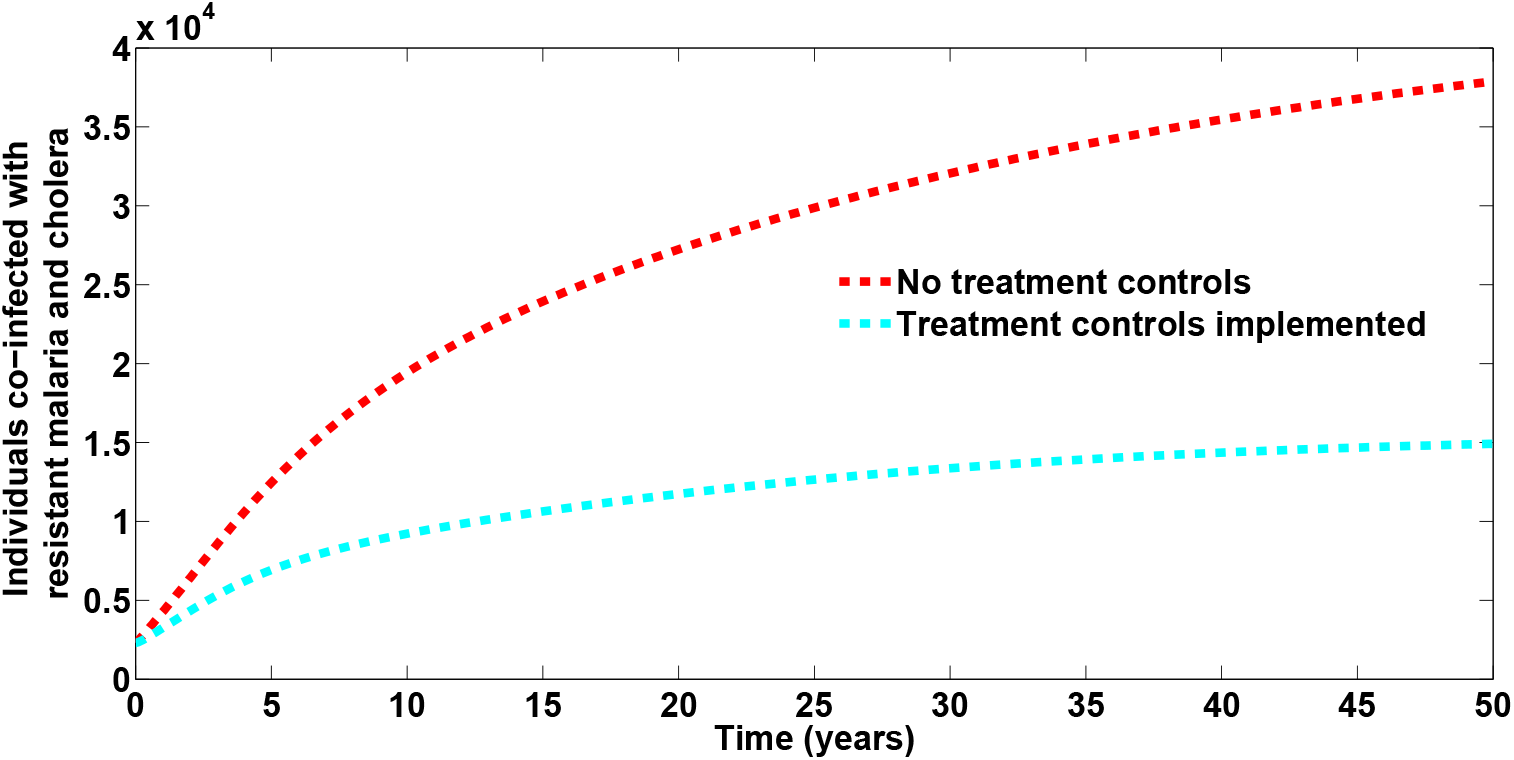
Simulations of the model (2) showing the total number of individuals co-infected with resistant malaria and cholera, when there is no drug resistance. Here, *σ*_M1_ = *σ*_M2_ = 0. All other parameters as in Table 2

**Figure 7:**
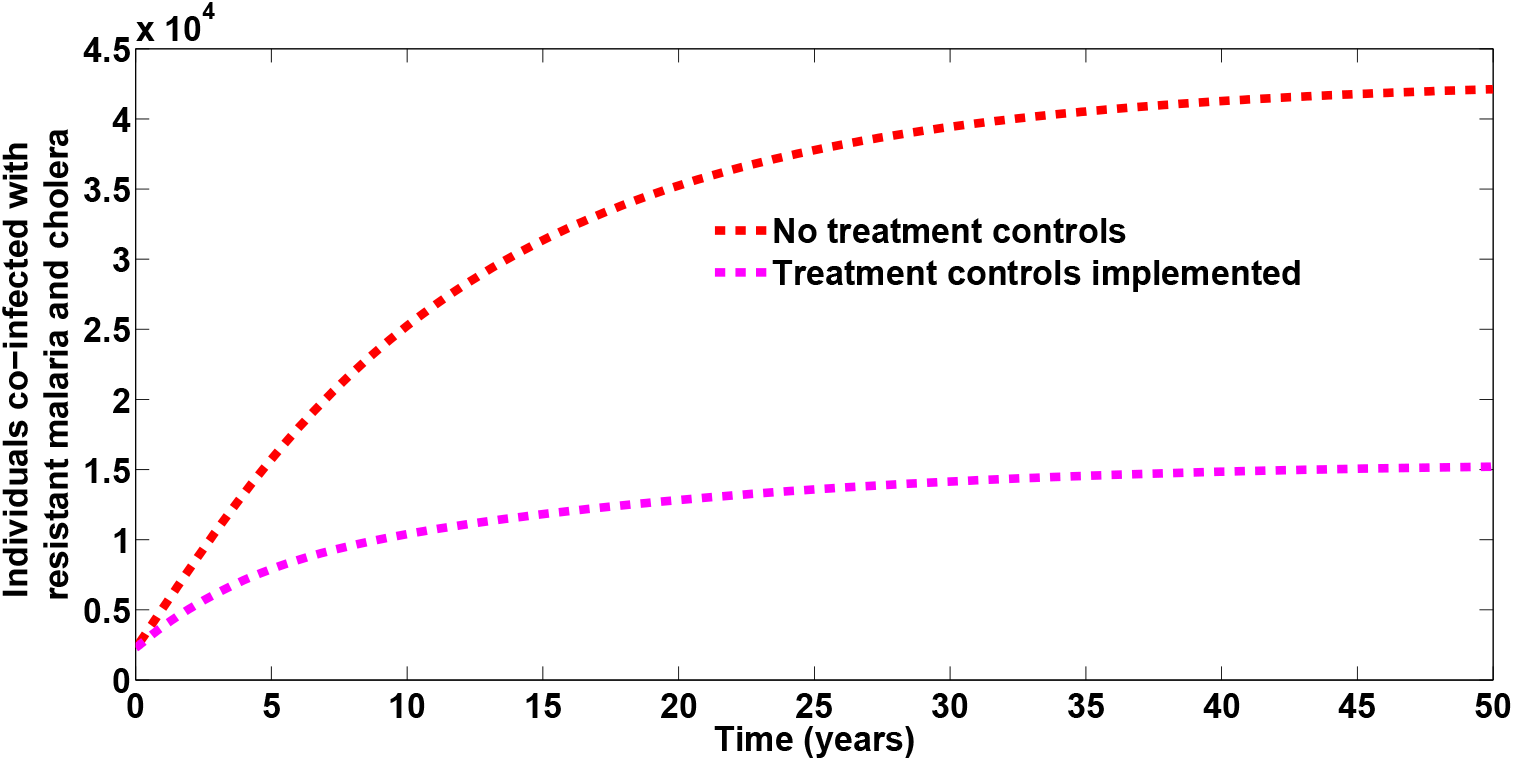
Simulations of the model (2) showing the total number of individuals co-infected with resistant malaria and cholera, when there is drug resistance. Here, *σ*_M1_ = *σ*_M2_ = 0.4. All other parameters as in Table 2

**Figure 8:**
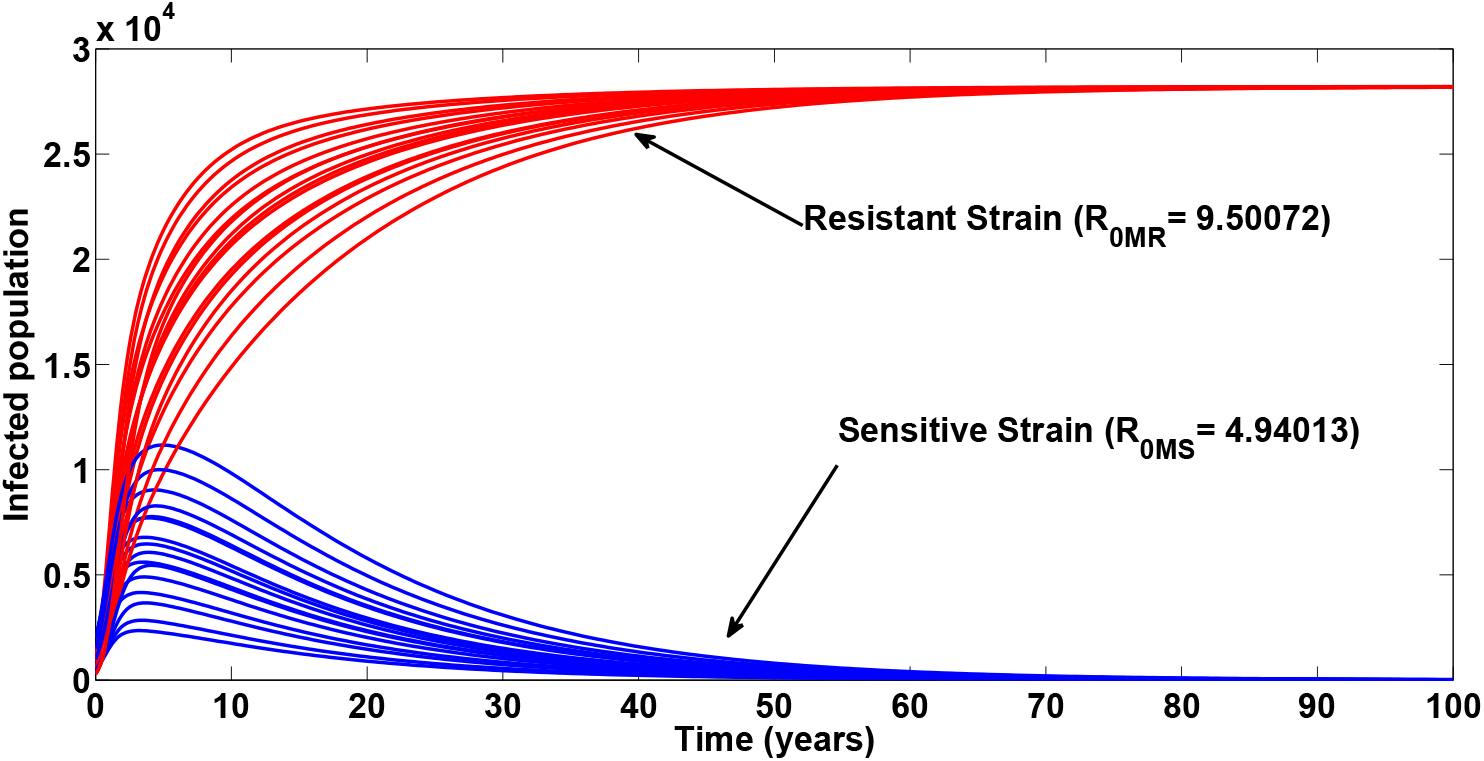
Simulations of the model (2) showing the total number of infected individuals at different initial conditions. Here, *β*_M_ = 7.034, *β*_7_ = 7.09, η_R_= 1.2, (so that 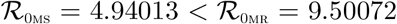). All other parameters as in Table 2

**Figure 9:**
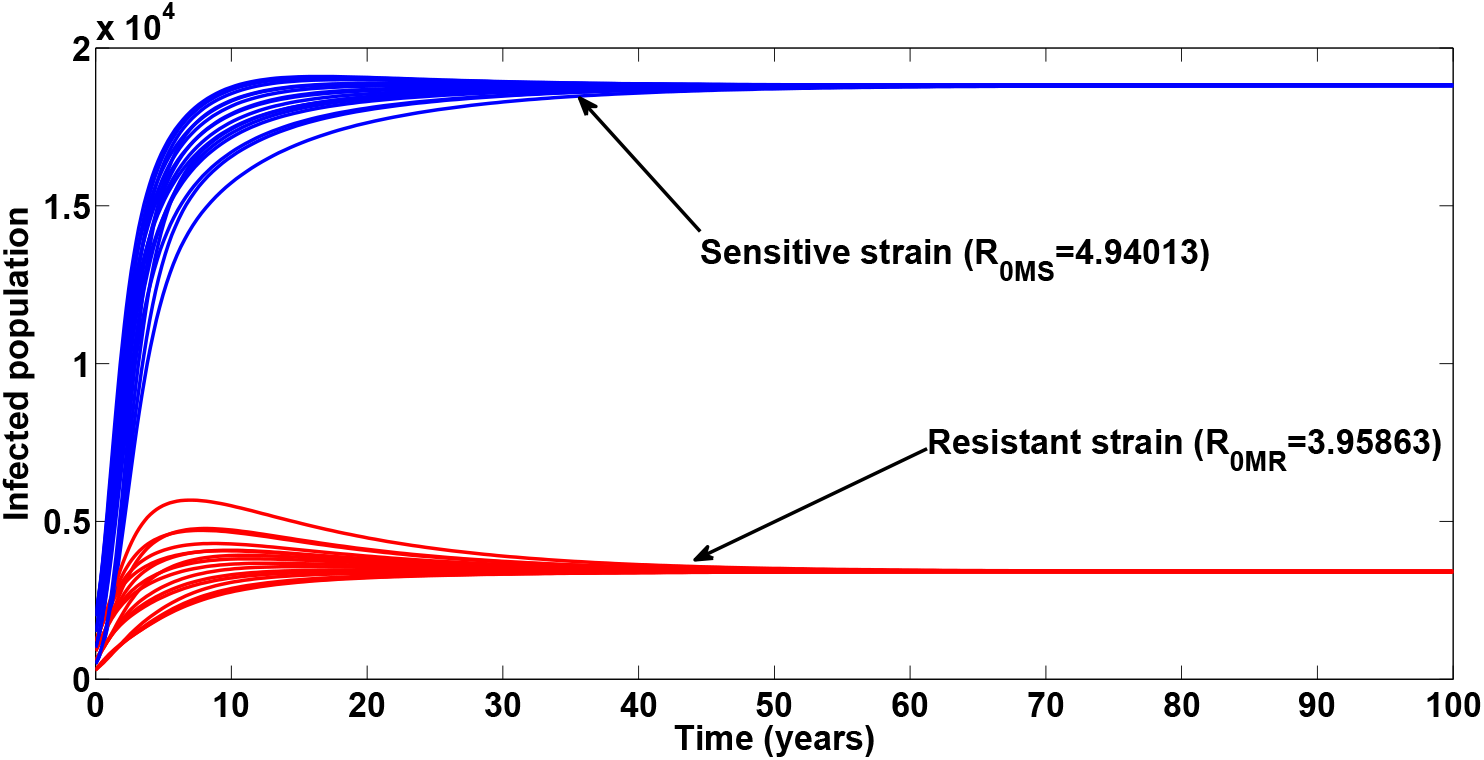
Simulations of the model (2) showing the total number of infected individuals at different initial conditions. Here, *β*_M_ = 7.034, *β*_7_ = 7.09, η_R_= 0.5, (so that 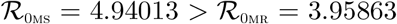). All other parameters as in Table 2

**Figure 10:**
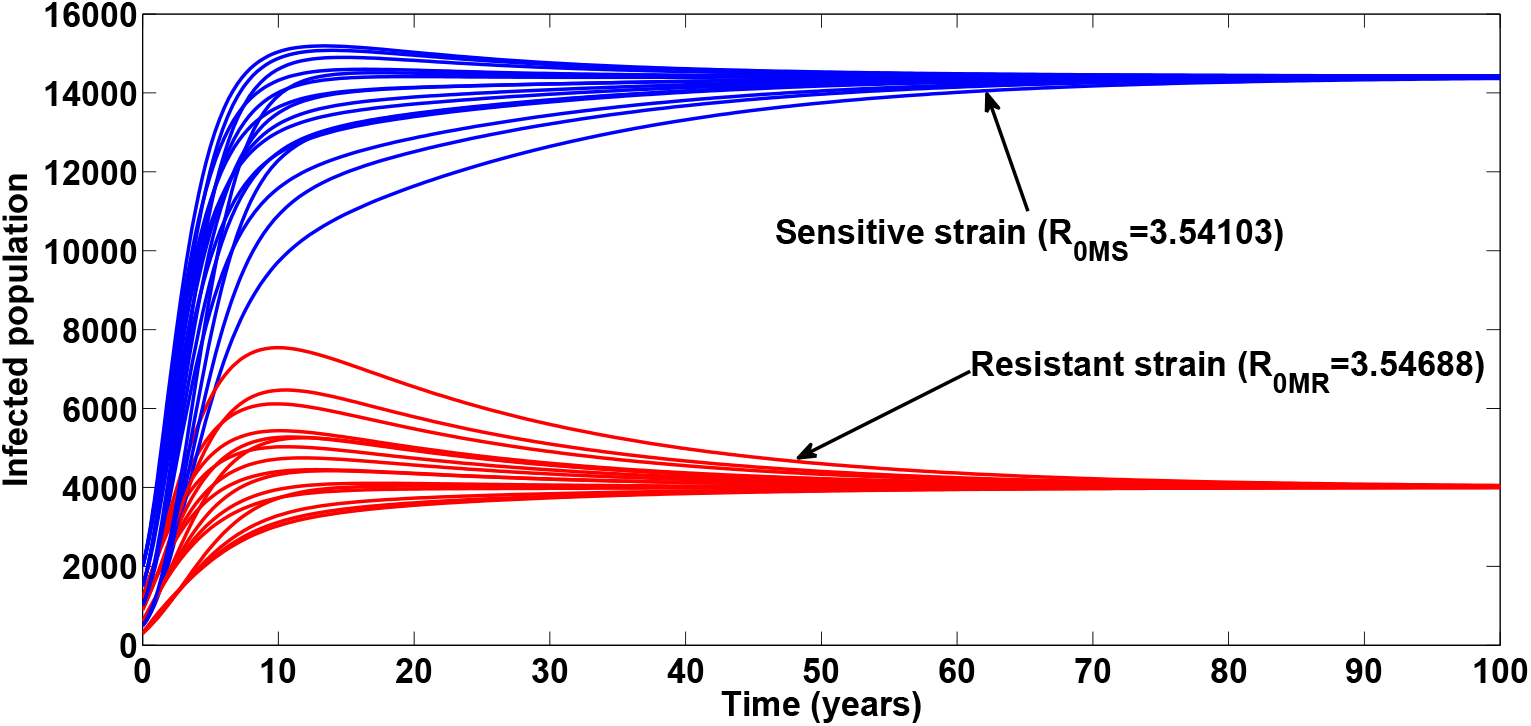
Simulations of the model (2) showing the total number of infected individuals at different initial conditions. Here, *β*_M_ = 5.034, *β*_V_ = 5.09, *η*_R_ = 0.625, (so that 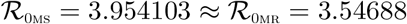). All other parameters as in Table 2

Figure 8 shows the simulation of the total number of infected individual at different initial conditions. It is observed that the resistant strain drives the sensitive strain to extinction when both reproduction numbers are greater than unity, with 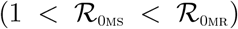. both strains co-exist with the sentitive strains dominating the resistaant strain when both reproduction number are greater than umity 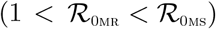. Likewise when both reproduction numbers are greater than unity with 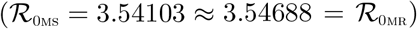,both strain co-exist with th sensitive strain dominating but not driving the resistant strain to extinction.

## 6 Conclusion

In this work, we have considered and analyzed a mathematical model for two strains of Malaria and Cholera with optimal control. The model assessed the impact of treatment controls in reducing the burden of the two diseases in a population, in the presence of malaria drug resistance. The model was shown to exhibit the dynamical property of backward bifurcation when the associated reproduction number is less than unity. The global asymptotic stability of the disease-free equilibrium of the model was proven not to exist. The necessary conditions for the existence of optimal control and the optimality system for the model is established using the Pontryagin’s Maximum Principle. Numerical simulations of the optimal control model revealed that malaria drug resistance can greatly influence the co-infection cases averted, even in the presence of treatment controls for co-infected individuals

## Data Availability

Not applicable

https://www.who.int/malaria/publications/

